# Understanding malnutrition management through a socioecological lens: Evaluation of a community-based child malnutrition program in rural Uganda

**DOI:** 10.1101/2021.09.01.21262681

**Authors:** Marie-Catherine Gagnon-Dufresne, Geneviève Fortin, Kirsten Bunkeddeko, Charles Kalumuna, Kate Zinszer

## Abstract

In Uganda, almost half of children under 5 years old suffer from undernutrition. Undernutrition, a common form of malnutrition in children, encompasses stunting, wasting and underweight. Causes of child undernutrition are complex, suggesting that interventions to tackle malnutrition must be multifaceted. In addition, limited access to healthcare for vulnerable populations restricts the potential of hospital-based strategies. Community-based management of acute malnutrition (CMAM), which includes nutritional counselling, ready-to-use therapeutic foods and the outpatient management of malnutrition by caregivers, is recognised as an effective approach for children’s recovery. However, evaluations of CMAM programs are largely based on biomedical and behavioural health models, failing to incorporate structural factors that influence malnutrition management. The objective of this evaluation was to understand the factors influencing malnutrition management in a CMAM program in rural Uganda, using the socioecological model to assess the multilevel determinants of outpatient malnutrition management. This evaluation used qualitative methods to identify determinants related to caregivers, healthcare and societal structures influencing children’s outpatient care. Data were collected at a community health clinic in 2019 through observations and interviews with caregivers of malnourished children. We observed 14 caregiver-provider encounters and interviewed 15 caregivers to examine factors hindering outpatient malnutrition management. Data were thematically analysed informed by the socioecological model. Findings showed that caregivers had a limited understanding of malnutrition. Counselling offered to caregivers was inconsistent and insufficient. Gender inequality and poverty limited caregivers’ access to healthcare and their ability to care for their children. Factors at the caregiver- and healthcare-levels interacted with structural factors to shape malnutrition management. Results suggest that CMAM programs would benefit from providing holistic interventions to tackle the structural barriers to children’s care. Using a socioecological approach to program evaluation could help move beyond individual determinants to address the social dynamics shaping malnutrition management in low- and middle-income countries.

**WHAT IS KNOWN?:**

- CMAM is recognised as a promising strategy to address moderate and severe acute malnutrition in children under five years old in resource-poor settings.
- Evaluations of CMAM programs largely focus on factors related to caregivers’ choices, behaviours, and practices to explain why CMAM has inconsistent results.
- Limited attention has been given to the multilevel determinants that influence the outpatient management of malnutrition in CMAM programs.

**WHAT THIS PAPER ADDS?:**

- Caregivers had limited understandings of malnutrition and its underlying mechanisms.
- Counselling provided to caregivers by program personnel was inconsistent and insufficient, often including contradicting information about treatments prescribed to children.
- Structural factors limited caregivers’ access to healthcare and their ability to comply with CMAM outpatient protocols.

## 1. INTRODUCTION

The United Nations Sustainable Development Goals call to ‘end hunger, achieve food security and improved nutrition, and promote sustainable agriculture’ by 2030 (United Nations, 2021). Yet, we are not on track to achieve this goal in less than a decade (United Nations, 2021). Nearly half of all deaths of children under 5 years of age are attributable to undernutrition globally, with low- and middle-income countries bearing most of the burden (World Health Organization, 2020). Undernutrition, a common form of malnutrition in children, includes chronic malnutrition or stunting (low height for age), acute malnutrition or wasting (low weight for height), as well as underweight (low weight for age) (Brown et al., 2009). In Uganda, child undernutrition is recognised as a public health priority and is a central pillar in various national policies (Franklin, 2020; National Planning Authority, 2013, 2020). Although some progress has been made in the country in recent years, it is estimated that 29% of children under five years old are stunted, 4% are wasted, and 11% are underweight (Uganda Bureau of Statistics & ICF, 2018). Causes of malnutrition in Uganda, including undernutrition, are complex. The Ugandan population is largely rural (84%) and nearly two thirds work in agriculture, making rural households vulnerable to environmental fluctuations like droughts and flooding, increasing food insecurity (USAID, 2021). With 35% of the population living in extreme poverty, access to sanitation and healthcare is limited (Mejia-Mantilla, 2020; USAID, 2021). High fertility rates and women’s limited access to employment also worsen food insecurity and malnutrition. Other drivers of child malnutrition include high disease burdens and poor feeding practices (USAID, 2021). This suggests that interventions tackling malnutrition must be multifaceted, and that limited access to healthcare for vulnerable populations restricts the potential of hospital-based strategies.

The World Health Organization (WHO) recognises community-based management of acute malnutrition (CMAM) as a promising strategy to address moderate and severe acute malnutrition in children under five years old in resource-poor settings, prompting its implementation in more than 60 countries (Reed & Kouam, 2013). CMAM includes community outreach, outpatient management of moderate acute malnutrition (MAM) and severe acute malnutrition (SAM) without complications, provision of ready-to-use therapeutic food (RUTF), and nutritional counselling (Shah More et al., 2018). The goal is to decentralise malnutrition management so most cases are treated at home by caregivers (Reed & Kouam, 2013).

Evaluations of CMAM programs are inconsistent, assessing a wide array of factors to explain why children’s outpatient care is suboptimal (Akparibo et al., 2017; Majamanda et al., 2014). These evaluations largely adopt behavioural or biomedical approaches to study malnutrition management, emphasising individual factors such as caregivers’ choices, behaviours, and practices. Factors examined notably include sharing or selling RUTF (Ashworth, 2006; Cohuet et al., 2012; Tadesse et al., 2015), giving complementary feedings in addition to RUTF (Ashworth, 2006; Kajjura et al., 2019), and treatment compliance (Atnafe et al., 2019; Yebyo et al., 2013). While these factors do influence malnutrition management, it would also be beneficial to assess determinants beyond the individual in CMAM program evaluations (Birn et al., 2017). Examining the multilevel determinants of children’s care in CMAM programs would highlight how caregivers’ behaviours are influenced by determinants at the healthcare- and structural- levels, limiting their ability to meet the long-term nutritional needs of their children.

The socioecological model is a useful framework for assessing malnutrition management holistically. Recognising that individuals are embedded in various interdependent environments (*i*.*e*., individual, interpersonal, community, organisational, policy), the socioecological model can be adapted and changed to suit research on various topics, including malnutrition (Lanfer & Reifegerste, 2021; Mahmudiono et al., 2019). It has proven useful for examining the complex interactions of individual and contextual factors in impacting health behaviours and outcomes (Tesfay et al., 2021).

In 2019, we conducted a qualitative evaluation of a CMAM program delivered by a community health clinic in rural Uganda to understand the various factors hindering the outpatient management of acute child malnutrition. We used the socioecological model to discuss three levels of determinants that could explain malnutrition management in this program, namely determinants linked to caregivers, healthcare, and societal structures. This article presents the results of this evaluation.

### 1.1 Program description

Located in the Eastern region of Uganda, Soft Power Health (SPH) is a not-for-profit organisation providing health education and affordable primary healthcare to rural, underserved communities through a community health clinic and community outreach programs (Soft Power Health, 2020). SPH’s community health clinic runs a program to treat children with MAM and SAM, which integrates all the key elements of CMAM programs as defined by the WHO (Reed & Kouam, 2013). SPH organises weekly community outreach activities in villages around the clinic. These activities involve malnutrition education and allow to screen for malnutrition in children of the participants present. There is also an on-site program offering nutritional counselling to caregivers of malnourished children at the clinic and provides high energy milk (HEM), a homemade mix of powdered milk, sugar, and oil, as treatment for malnutrition. High in calories and protein, HEM is cheaper to make than commercial RUTF and can be easily reproduced by caregivers at home if needed (Namayengo, 2001).

When coming to the clinic, children go through three program stages. First, children’s anthropometric measurements are taken at the triage station. Second, children and their caregivers are sent for a consultation with a doctor for confirmation of diagnosis or follow-up. The doctor also prescribes HEM according to children’s age and weight. Third, the kitchen personnel distribute HEM and explain to caregivers how to prepare the treatment. Program personnel include triage officers, doctors, and support staff. Nutritional counselling should be provided at each stage by corresponding personnel. Children are expected to return to the clinic every two weeks for re-evaluation of their nutritional status. They are followed until their anthropometric measurements indicate recovery. In 2019, it was estimated that more than 6,200 paediatric patients were screened for malnutrition at the clinic and that 500 received HEM as treatment for malnutrition (Soft Power Health, 2019).

## 2. MATERIALS & METHODS

### 2.1 Evaluation design

We conducted a qualitative process evaluation of SPH’s on-site malnutrition program from September to December 2019. The evaluation responded to the need of program managers to better understand factors that could hinder children’s nutritional recovery, with the prospect of improving program services. This evaluation focused on the encounters between caregivers of malnourished children enrolled in the program and program personnel to examine caregivers’ knowledge of malnutrition and perceptions of the program, program delivery, as well as contextual factors influencing the outpatient management of malnutrition (Moore et al., 2015). Data were collected through non-participant observations of patient-provider encounters during program delivery at the clinic, and subsequent semi-structured interviews with caregivers.

### 2.2 Sampling strategy

We used purposive sampling to recruit caregivers of children between 12 and 59 months old diagnosed with MAM or SAM, enrolled in SPH’s malnutrition program, and receiving HEM as treatment. The WHO criteria for diagnosis of child MAM and SAM, also used by the clinic, were adopted as inclusion criteria (World Health Organization, 2006). Exclusion criteria were as follows: 1) children under 1 year old, as they receive different treatments for malnutrition; 2) children over five years old, as diagnosis criteria for malnutrition differ; and 3) children with cerebral palsy, as they are prescribed HEM as long-term dietary supplement. During the 18 days of data collection in October and November 2019, all caregivers of eligible children were approached at the triage station, and all accepted to participate (N=15). All gave their oral consent for observation of their encounters with program personnel and participation in an interview. Before the interviews, caregivers gave their written informed consent in a form written in Lusoga (local language), which was read for them if needed. Program personnel involved in the consultations also provided their oral consent for the observations.

### 2.3 Data collection and translation

We observed a total of 14 encounters between caregivers and program personnel and interviewed 15 caregivers (two caregivers sharing custody of the same child were observed together but interviewed separately). Observations were conducted directly after recruitment to ensure that all three stages of the program could be observed. Observations allowed to examine the encounters between caregivers and program personnel to evaluate factors associated with program delivery and counselling that could influence children’s care (Liu & Maitlis, 2010).

Immediately after observation of patient-provider encounters and counselling, semi-structured interviews were conducted in Lusoga and English with the same caregivers. Interviews assessed factors linked to caregivers’ practices, knowledge, and living conditions that could influence outpatient malnutrition management. We also examined their perceptions of the program, including nutritional counselling and HEM preparation. At the end of each interview, caregivers received a pictogram tailored to their children’s HEM treatment (*i*.*e*., quantity and intervals between feeds) explaining how to prepare it. If unable to describe how to prepare HEM, caregivers were referred to program personnel for further counselling. Interviews, lasting between 12 and 29 minutes, were conducted in a quiet room away from the main clinic.

Lusoga was the mother tongue of all participants. Therefore, observations and in-depth interviews were conducted in collaboration with a local interpreter, previously trained by the first author. The interpreter provided whisper translation of the exchanges between program personnel and caregivers during observations and translated all interviews in real time. The interpreter validated her initial translations by adjusting interview transcripts while listening to the recorded interviews, increasing translation precision and validity (Shimpuku & Norr, 2012; Squires, 2009).

### 2.4 Data analysis

This study used interpretive description to analyse interview and observation data, allowing for a contextually situated analysis of children’s care which could be useful for improving program practices (Thorne et al., 1997, 2004). Data were transcribed in English, imported in NVivo, and submitted to thematic analysis (Braun & Clarke, 2006). Analysis was done in a deductive and inductive manner to examine determinants of care from a socioecological perspective, while allowing for developing codes directly from the data (Lanfer & Reifegerste, 2021; Saldaña, 2013). Notes from observations were used to validate analysis of interview data. Themes were confronted to literature on the evaluation of CMAM programs in a logic of data triangulation between observations, interviews and existing literature (Nowell et al., 2017).

### 2.5 Ethical considerations

Ethics approval was granted by Mbarara University of Science and Technology’s Research Ethics Committee in Uganda. An exemption was granted after evaluation of the project by the University of Montréal’s Ethics Committee for Science and Health Research following Article 2.5 of the Panel on Research Ethics of the Government of Canada. This project was conducted in an ethical manner. Informed consent was obtained from participants. Data were anonymised to protect participants’ confidentiality.

## 3. RESULTS

We observed 14 caregiver-provider encounters and interviewed 15 caregivers of malnourished children. Sociodemographic characteristics of participants are presented in Table 1. Three themes were developed from the analysis of the 15 interviews. The first theme presents determinants related to caregivers’ knowledge and practices that may influence the outpatient management of malnutrition. The second theme identifies healthcare determinants that can impact how caregivers comply to treatment protocols at home. The third theme describes the structural determinants that may affect children’s care. Notes from program observations were used to enrich and validate findings from interview data. Summarised observations are represented in Table 2.

**Table 1.** Sociodemographic characteristics of participants.

| **Sociodemographic characteristics** | ***n*** | **% or mean (range)** |
| --- | --- | --- |
| **Sex of caregivers** |  |  |
| Women | 13 | 87% |
| Men | 2 | 14% |
| **Relationship of caregivers to children** |  |  |
| Mother | 9 | 60% |
| Father | 2 | 13% |
| Grandmother | 3 | 20% |
| Aunt | 1 | 7% |
| **Age of caregivers** |  |  |
| Age in years: mean (range) | - | 33 (21-56) |
| **Marital status** |  |  |
| Married | 8 | 53% |
| Married, not living with partner | 3 | 20% |
| Unmarried | 4 | 27% |
| **Self-reported religious affiliation** |  |  |
| Muslim | 5 | 33% |
| Catholic | 3 | 20% |
| Anglican | 4 | 27% |
| Born-again Christian | 3 | 20% |
| **Highest school grade completed** |  |  |
| Grade: mean (range) | - | 6<sup>th</sup> (none-12<sup>th</sup>) |
| **Women employed** |  |  |
| Number of women employed | 4 | 31% |
| **Cost of transportation between home and clinic, one way in Ugandan shillings: mean (range)** |  |  |
| Cost in USD: mean (range) | - | \$2.13 (\$0.43-\$4.25) |
| **Sex of children** |  |  |
| Girls | 6 | 38% |
| Boys | 10 | 62% |
| **Age of children** |  |  |
| Age in months: mean (range) | - | 24.5 (12-51) |
| **Diagnosis** |  |  |
| Moderate acute malnutrition | 11 | 60% |
| Severe acute malnutrition | 5 | 31% |
| **Duration of HEM\* treatment** |  |  |
| Duration in months: mean (range) | - | 2 (0-12) |
| **Number of children in the family** |  |  |
| Number of children: mean (range) | - | 3 (1-8) |

\*HEM: high energy milk, a homemade mix of powdered milk, sugar, and oil, used as treatment for malnutrition in Soft Power Health's program, replacing RUTF as usual treatment in CMAM.

**Table 2.** Summarised observations of SPH’s CMAM program delivery.

| Stage of the program and elements assessed | Summarised results |
| --- | --- |
| **Triage station (triage officers)**<br>• Recording of all anthropometric measurements<br>• Information on the extent and content of counselling provided (if any)<br>• Time allotted for questions by program personnel, questions asked by caregivers and answers provided<br>• Knowledge of malnutrition demonstrated by program personnel<br>• Comments heard and other observations | • Anthropometric measurements were taken systematically for all children observed.<br>• Around one third of caregivers received no counselling at the triage station.<br>• Information shared during counselling varied by program personnel.<br>• Counselling mostly focused on how to have a balanced diet, seldom addressed the signs of malnutrition.<br>• Most of the time, program personnel did not ask caregivers if they had questions about their children's condition.<br>• Counselling was often skipped if the clinic was very busy or when the most experienced program members were absent.<br>• Program personnel mentioned not counselling caregivers if they remembered that they were counselled in previous visits.<br>• When counselling was provided, triage personnel showed a sufficient understanding of the causes and consequences of malnutrition.<br>• Personnel was supportive of caregivers and helped find solutions when families faced challenges that could hinder children's care. |
| **Medical consultation (doctors)**<br>• Validation of anthropometric measurements from patient card<br>• Physical examination to look for visible signs of malnutrition and discussion with caregivers<br>• Information on the extent and content of counselling provided (if any)<br>• Time allotted for questions by doctors, questions asked by caregivers and answers provided<br>• Knowledge of malnutrition demonstrated by doctors<br>• Prescription of HEM, calculation of formula distribution using the WHO chart, explanation of treatment plan to caregivers<br>• Comments heard and other | • Signs of malnutrition, including anthropometric measurements, were assessed for most children observed.<br>• Counselling was provided to most caregivers, and mostly focused on how to have a balanced diet. Counselling seldom addressed the signs of malnutrition.<br>• Doctors only explained the HEM treatment plan (*i.e.*, amount of HEM and interval between feeds) prescribed to malnourished children to two caregivers.<br>• Doctors often asked caregivers if they had questions about their children's conditions.<br>• Less experienced doctors provided very little counselling and did not calculate the amount of HEM prescribed to children using the WHO chart.<br>• Two women doctors discussed the impacts of sociocultural contexts on malnutrition, including family planning and women's access to |
| observations | employment. These issues were not addressed by men doctors.<br>• Doctors mentioned that most of the counselling should be provided at other stages of the program, as doctors have a busy schedule. |
| **Distribution of HEM (support staff)**<br>• Preparation of HEM formula with caregivers<br>• Explanation of HEM preparation and treatment plan to caregivers<br>• Information on the extent and content of counselling provided (if any)<br>• Time allotted for questions by program personnel, questions asked by caregivers and answers provided<br>• Knowledge of malnutrition demonstrated by program personnel<br>• Comments heard and other observations | • The HEM treatment was never prepared in front of caregivers.<br>• Program personnel explained how to prepare HEM and the treatment plan (*i.e.*, amount of HEM and interval between feeds) to less than half of caregivers.<br>• The treatment plan discussed with caregivers sometimes contradicted that prescribed by doctors in the previous program stage.<br>• Counselling, which was provided to around two thirds of caregivers, mostly focused on how to have a balanced diet.<br>• Other counselling provided addressed domestic violence, the importance of sleeping under a mosquito net, and how to clean dishes properly.<br>• When counselling was provided, personnel showed sufficient understanding of the causes and consequences of malnutrition.<br>• Personnel often asked caregivers if they had questions about their children's conditions or the HEM treatment.<br>• Personnel mentioned that caregivers of children who had been enrolled in the program for some time did not require as much counselling. |

### 3.1 Lacking the knowledge to support their children: Determinants of malnutrition management at the caregiver-level

#### 3.1.1 Having a limited understanding of malnutrition

When asked about their understanding of malnutrition, caregivers explained what it meant to them and discussed signs of child malnutrition. The primary sign identified was weight loss, as many caregivers said that losing weight led to seeing the children’s bones under their skin. Most described malnutrition as “thinning out” or “growing small.” Accordingly, a woman in her 50s (P05) explained how she understood that her child was malnourished: “The signs that showed me that she had gotten malnourished were that I saw her thinning out, with no flesh onto the bones.” A woman in her 20s (P07) defined malnutrition similarly: “There are some kids in this case of malnutrition, you can find that they have gone small, and they have no flesh. You just see bones.” Although caregivers interviewed could generally identify weight loss as a sign of malnutrition, many of their children had symptoms other than weight loss. Only a few participants named other signs of malnutrition, such as “swelling of the cheeks,” “enlarging of the abdomen,” “yellowing of the face” or “a change of colour of the hair.” Caregivers often asked for more information on malnutrition, suggesting that their health literacy was limited and that they would have benefitted from better understanding this condition.

#### 3.1.2 Not grasping the mechanisms that bring malnutrition

Caregivers discussed causes of malnutrition and factors that contribute to improving children’s nutritional status. Caregivers named many perceived causes of malnutrition that were not necessarily linked with nutrition such as “sleeping well,” “giving leftover food,” “the body being hot,” “coldness,” and “delaying seeking medical care.” Nevertheless, all but one participant identified inappropriate feeding habits as the primary cause of malnutrition. A woman in her 50s (P08) explained that not eating nutritious food could cause malnutrition: “What brings malnutrition: this child not feeding well or not eating food which can help build the body.” A woman in her 20s (P02) described how she could help her children recover from malnutrition: “According to me, the way I think, it means giving enough food to this child and having different meals, like having a balanced diet.” Although all participants stressed their role as caregivers in supporting their children to recover from malnutrition, the majority mentioned mostly feeding their children on a carbohydrate diet. Posho (cornmeal porridge), matooke (boiled green banana), rice and potatoes were the foods most named by caregivers: all participants named at least one of these food items and almost half only discussed carbohydrates. For instance, a woman in her 50s (P09) detailed her child’s daily diet: “There are many foods, like posho, potatoes, matooke. Sometimes, the father also brings in rice.” Data suggests that while caregivers could identify perceived causes of malnutrition, they did not fully grasp the mechanisms of malnutrition or know concrete ways to support their children.

### 3.2 Being unequipped to make informed decisions: Healthcare determinants of malnutrition management

#### 3.2.1 Difficult knowledge transfer and inconsistent counselling

The counselling provided to caregivers by program personnel varied greatly. This was assessed during observations of patient-provider encounters and confirmed during interviews with caregivers. The frequency, content, and intensity of counselling were inconsistent across program stages and personnel. Counselling sessions most often addressed how to have a balanced diet, when to feed children, and examples of local foods to give them. A woman in her 20s (P12) confirmed this emphasis put on diet when remembering counselling: “I was told to cook food […] Taking care, giving the treatment that they have given me, and also giving food all the time.” However, others mentioned receiving very little or no counselling. Another woman in her 20s (P01) explained that in her previous appointment, when her children were first admitted into the program, she did not receive much counselling: “Last time when I came, I didn’t get enough education. It’s today that I have got more health education.” Similarly, another woman in her 20s (P07) who had previously dropped out of the program and returned after months of absence said that she was not counselled: “There is nobody who educated me […] They taught me how to cook only […] I also had the first teaching on the other side about hygiene.” Therefore, in line with our observations, counselling offered to caregivers was inconsistent. Knowledge transfer also seemed inefficient, as caregivers did not all realise that they had received counselling, nor did they fully understand the information that was shared with them. Many participants expressed their uncertainty during the interviews and asked for recommendations to better care for their children.

#### 3.2.2 Confusion caused by conflicting information on the HEM treatment

All participants had positive perceptions of HEM. Many mentioned that HEM was “better than cow’s milk,” “built the body” and “helped children gain energy.” However, most were hesitant when asked to describe the treatment. A woman in her 20s (P06) explained that she did not know what HEM was made of: “Ah! For me I don’t know, but they say they put that building food in the milk.” Likewise, a woman in her 40s (P04) expressed not understanding what was HEM: “This milk you are giving me? I don’t know it.” In addition to not knowing what HEM was made of, many participants were confused when asked to explain how to prepare HEM. As confirmed from program observations, this could be explained by the fact that caregivers were often given different information about the prescribed amount of HEM and feeding times by doctors and other personnel. For example, in several instances, instructions given by support staff upon distribution of HEM contradicted the treatment plan previously described by doctors. Considering that the treatment plan changed at every visit (*i.e*., according to children’s weight, height, and nutritional status), this might influence treatment compliance.

While some were given conflicting information about the HEM treatment, others received no information about how to prepare it, such as this woman in her 20s (P07) who expressed not being taught about how to cook HEM: “I wasn’t told […] How long does that milk take to boil? I forgot asking.” Similarly, a woman in her 50s (P08) was unsure about how to measure the right quantity of HEM for her child:

> This measuring part, like measuring milk… I was told to measure with the baby cups, the ones having measurements. What if I just give the estimated amount? Now, this one is a big child, what if I use a regular cup? […] What if I don’t use the real measurements?

At the end of their visit in the program, participants were therefore still confused about how to prepare HEM. Yet, few participants were asked about their understanding of the treatment plan by program personnel. Importantly, given their limited access to stable incomes and education, many participants did not own baby bottles with measurements, and did not know how to read, making it impossible for them to comply with the treatment prescribed. This was not addressed by program personnel, while most participants would have needed more information. Having asked participants about their understanding of the prescribed HEM treatment and its preparation could potentially have improved compliance.

### 3.3 Trying to overcome inequalities: Structural determinants of malnutrition management

#### 3.3.1 Poverty hindering access to healthcare and compliance with CMAM protocols

Many caregivers named poverty as an obstacle to managing their children’s condition. One way in which economic insecurity impacted malnutrition was by restricting access to healthcare. Most caregivers, like this woman in her 50s (P05), recognised access to medical care as crucial for nutritional recovery: “What will help her is taking the child to see the medical personnel […] and giving treatment so that the child will get fine.” However, many participants lived far from the clinic and had to pay up to 8.50 USD for a round trip using public transport, even if they had no income. For instance, a woman in her 20s (P01) explained delaying to seek medical attention for her child, not having enough money to pay for transport: “When he feels sick, you may lack money for transport to bring the child to the clinic. Like last time, I was supposed to come back on [a specific day] but money was a problem.” Another woman in her 20s (P02) who had been receiving free healthcare and HEM from the program expressed being scared because program personnel told her she would soon have to start paying for treatment:

> Right now, since I’m not working, I don’t have money. And now, they are saying that they are going to stop [my children] from getting milk […] I don’t have any money, and I don’t have anywhere to start from to get money, and I’ve been relying on this milk.

Another caregiver (P05) mentioned that poverty hindered her ability to comply with the recommendations of the program, lacking the money to buy a variety of food to feed her child with a balanced diet: “Maybe I can [help my child get better], but right now I don’t have enough money. Still, I can buy what I can afford.” Consequently, poverty limited access to healthcare and hindered treatment compliance for participants, as they often could not afford to pay for transport, treatment, or diverse foods to support their children.

#### 3.3.2 Gender inequality limiting caregivers’ ability to care for malnourished children

Out of 15 participants interviewed, 13 were women. Most of them did not access education beyond primary school and were not employed. In contrast, the two male respondents had secondary education and were expected to support their family financially. Women participants were primary caregivers, responsible for much of child rearing duties, often having many children to care for. They mentioned that difficult family situations impacted their ability to manage their children’s malnutrition. For instance, a woman in her 30s (P15) with many children explained that people in her village provided food for them because she did not have an income. She said that her husband, who had abandoned her and their children, occasionally came back to force her to have other children, while not supporting his family:

The man is not at home. He has taken a long time without coming home. He ran away from home, and he has spent years [without coming back] […]. He just forced me to have sex. Then, I gave birth to this one.

She also expressed that while many of her children were malnourished, she did not have money to bring them all to the clinic.

Other women similarly discussed being left to provide for their children alone. One woman (P01) said that she moved out of her husband’s home because he “did not give [her] care” and that her financial situation was “very bad.” Another woman (P12) mentioned that her husband was rarely present: “I stay with him, but still… I stay with him for a short time. […] He’s having two wives so he’s not ever at home.” Similarly, a woman in her 20s (P02) discussed seeking help from the clinic’s domestic violence counsellor to ensure she could provide treatment for her children: “I went to the [domestic violence] counsellor who came and spoke to [the program personnel], and they made for me a receipt indicating that I will be taking milk for free, and also the transport included.” Therefore, difficult family situations and women’s financial dependency upon their male partners seemed to hinder their ability to respond to their children’s nutritional needs and access services required to overcome malnutrition.

## 4. DISCUSSION

This article offers novel insight on the outpatient management of child malnutrition in the context of CMAM, following the evaluation of a malnutrition program in rural Uganda. It provides more information on the multifaceted determinants of malnourished children’s care. Three themes were developed, indicating that caregiver/household, healthcare, and structural determinants can shape how children fare in programs where acute malnutrition is to be treated by caregivers at home. To the best of our knowledge, this is the first CMAM program evaluation that has used the socioecological model as conceptual framework to guide its findings. Results should be used to inform the implementation and evaluation of these programs in low- and middle-income countries to ensure that children can thrive.

In our evaluation, we found that a majority of caregivers experienced multiple causes of marginalisation – including chronic poverty, gender inequalities, limited access to education and employment, and lack of social protections – which clustered to hinder children’s care (Nisbett & Harris, 2018). These structural barriers increased the vulnerabilities of caregivers and their families, limiting their agency to become food secure, access healthcare, and care for malnourished children (Clement et al., 2019). Yet, many evaluations of CMAM programs adopt biomedical or behavioural health approaches, neglecting to explore factors beyond the individual that can influence malnutrition management (Abubakar, 2013). It has been argued that the multilevel determinants of children’s health should be studied concurrently when assessing health interventions, to grasp how individual factors interact with structural factors to shape patterns of health vulnerability and inequity (Nisbett & Harris, 2018; Uddin et al., 2021). CMAM programs should therefore consider these barriers and examine how structural determinants can hinder caregivers’ capacity to uphold CMAM protocols (Kim et al., 2013).

The first theme demonstrated that caregivers’ characteristics and practices could influence malnutrition management. Our results are consistent with evaluations of other CMAM programs showing that caregivers in low-resource settings may have limited understandings of malnutrition (Isanaka et al., 2020; Mbogo et al., 2020; Peter et al., 2019; Ware, 2018). Our findings showed that most caregivers associated nutrition with weight loss, having more difficulty identifying other signs of malnutrition. Interestingly, a study investigating community perceptions of child undernutrition and CMAM in India found that caregivers were less likely to consult health professionals if their children were ‘only skinny’ (Burtscher & Burza, 2015). This result is worth considering in the Ugandan context, as it suggests that low health literacy regarding malnutrition could impede caregivers’ ability to detect the condition early, recognise its severity, and seek medical care. Caregivers’ perceptions of malnutrition could also influence treatment compliance and children’s care.

Our second theme identified determinants associated with program implementation that could shape the outpatient management of malnutrition. Observations and interviews showed that caregivers received inconsistent counselling, as well as insufficient (and sometimes contradicting) information about their children’s treatment plans. This resonates with findings from other CMAM evaluations highlighting that healthcare providers do not always adhere to program protocols, and that training and supervision of personnel are critical to programmatic success (Shah More et al., 2018; Tadesse et al., 2016). Studies also found that nutritional counselling was often weak in these programs (Ashworth, 2006; Somassè et al., 2016). These results are significant for our evaluation, potentially explaining why caregivers struggled to describe and comply with the treatment prescribed. Strengthening knowledge transfer in CMAM programs to ensure that caregivers better understand treatment plans could improve children’s care.

Our third theme outlined structural barriers impacting outpatient malnutrition management. Structural violence, referring to the social structures embedded in everyday practices and institutions that exacerbate health inequalities and exposure to risk for certain individuals, can limit the agency of caregivers in taking care of their malnourished children (Farmer et al., 2006; Nisbett & Harris, 2018; Chung & Hunt, 2012; Chung, 2021). In our evaluation, caregivers, who were mostly women, stressed that poverty was an obstacle to follow CMAM protocols, limiting their ability to pay for transport, seek medical care, and buy a diverse diet for children. Their lack of access to education and employment also increased their financial dependency upon their husbands and left them in precarious situations when unmarried.

While many studies on nutrition highlight that maternal education, poverty and gender inequality can influence children’s nutritional status (Kajjura et al., 2019; Kumeh et al., 2020; Nankinga et al., 2019; Uddin et al., 2021), few evaluations of CMAM programs consider these factors in their analysis. These studies recommend assessing economic and food needs of entire households, family dynamics, domestic violence, and women’s empowerment (Shah More et al., 2018; Sharma & Subramanyam, 2021; Tette et al., 2016). Assessing these structural determinants of care in program evaluations could help deconstruct the ‘gendered and classed discourses of blame’ often present in food-related research, in which women and their behaviours are blamed for family feeding practices (Mackay, 2019). Our results, supported by the existing literature, therefore suggest that CMAM programs should adopt integrated responses, tackling the acute needs of malnourished children while also intervening to improve the social conditions at the roots of children’s nutritional trajectories.

### 4.1 Limitations

First, it is possible that program personnel and caregivers adapted their behaviours as they were being observed and interviewed. This could have altered how counselling was provided by program personnel (*i.e*., leading to more thorough counselling). It could also have changed the ways in which caregivers responded to interview questions (*i.e*., saying they were satisfied with the CMAM program), despite being encouraged to openly share the positive and negative aspects of their experience. Second, working with an interpreter in cross-cultural research can impact the reliability and validity of findings, because subtle differences in meaning across languages and cultures are not always captured (Kapborg & Berterö, 2002). It is therefore possible that some sociocultural references were not grasped during data collection and analysis. However, training of the interpreter before conducting the interviews helped minimise these issues. This contributed to ensuring that interview transcripts conveyed conceptual equivalence, meaning that the interpreter translated not only the literal meanings of words, but also conveyed how words relate conceptually in their context (Squires, 2009). In addition, the fact that the first author was immersed in the evaluation context for a prolonged period is an important strength of this evaluation. Close collaboration with local healthcare personnel throughout the process also ensured that the content of the evaluation was ethical, transparent, and culturally appropriate.

## 5. CONCLUSION

CMAM programs could be improved by extending their scope beyond caregivers’ feeding practices and care behaviours. Programs should strengthen their counselling curricula to ensure that caregivers receive sufficient information on children’s nutritional care. Counselling should also be used to screen for structural vulnerabilities affecting households, to tackle the multilevel barriers to malnutrition management. CMAM programs being often delivered by non-governmental organisations providing primary healthcare, caregivers could be referred to partner services targeting chronic food insecurity, poverty, domestic violence, family planning, and women’s empowerment. We hope that the results from this evaluation contribute to encouraging CMAM programs to develop tailored responses to the needs of households, incorporating complex interventions to address structural determinants of care (Birn et al., 2017; Kim et al., 2013; Nisbett et al., 2014). As we step into the last decade of the Sustainable Development Goals, it is critical that CMAM programs involve multidisciplinary teams to support families in accessing the resources needed to tackle the fundamental causes of undernutrition.

## Data Availability

To protect the confidentiality and safety of evaluation participants, qualitative data from direct observations and semi-structured interviews are not available.

## ACKNOWLEDGEMENTS

We would like thank Sandra Nambi who facilitated the interview process by providing translation and who gave generous advice for the design and conduct of this evaluation. We would also like to thank Dr Jessie Stone and all the Soft Power Health personnel for their support, as well as evaluation participants who dedicated their time to participate in interviews. The first author received a Queen Elizabeth Scholarship from Universities Canada and a mobility award from the University of Montréal through the Ministry of Education and Higher Education of Québec to undertake a graduate project in Uganda.

## REFERENCES

Abubakar, A. (2013). Psychosocial Aspects of Malnutrition Among African Children: Antecedents, Consequences, and Interventions. In M. J. Boivin & B. Giordani (Eds.), Neuropsychology of Children in Africa: Perspectives on Risk and Resilience (pp. 181–202). Springer. https://doi.org/10.1007/978-1-4614-6834-9_9

Akparibo, R., Lee, A., & Booth, A. (2017). Recovery, Relapse, and Episodes of Default in the Management of Acute Malnutrition in Children in Humanitarian Emergencies: A systematic review. Oxfam. https://doi.org/10.21201/2017.9149

Ashworth, A. (2006). Efficacy and Effectiveness of Community-Based Treatment of Severe Malnutrition. Food and Nutrition Bulletin, 27(3_suppl3), S24–S48. https://doi.org/10.1177/15648265060273S303

Atnafe, B., Roba, K. T., & Dingeta, T. (2019). Time of recovery and associated factors of children with severe acute malnutrition treated at outpatient therapeutic feeding program in Dire Dawa, Eastern Ethiopia. PLOS ONE, 14(6), e0217344. https://doi.org/10.1371/journal.pone.0217344

Birn, A.-E., Pillay, Y., & Holtz, T. H. (2017). Textbook of Global Health (Fourth edition). Oxford University Press. https://doi.org/10.1093/acprof:oso/9780199392285.001.0001

Braun, V., & Clarke, V. (2006). Using thematic analysis in psychology. Qualitative Research in Psychology, 3(2), 77–101. https://doi.org/10.1191/1478088706qp063oa

Brown, K. H., Nyirandutiye, D. H., & Jungjohann, S. (2009). Management of children with acute malnutrition in resource-poor settings. Nature Reviews Endocrinology, 5(11), 597–603. https://doi.org/10.1038/nrendo.2009.194

Burtscher, D., & Burza, S. (2015). Health-seeking behaviour and community perceptions of childhood undernutrition and a community management of acute malnutrition (CMAM) programme in rural Bihar, India: A qualitative study. Public Health Nutrition, 18(17), 3234– 3243. https://doi.org/10.1017/S1368980015000440

Chung, R. (2021). Structural health vulnerability: Health inequalities, structural and epistemic injustice. Journal of Social Philosophy, 52(2), 201–216. https://doi.org/10.1111/josp.12393

Chung, R., & Hunt, M. R. (2012). Justice and Health Inequalities in Humanitarian Crises: Structured Health Vulnerabilities and Natural Disasters1. In Health Inequalities and Global Justice. Edinburgh University Press. https://doi.org/10.3366/edinburgh/9780748646920.003.0012

Clement, F., Buisson, M.-C., Leder, S., Balasubramanya, S., Saikia, P., Bastakoti, R., Karki, E., & van Koppen, B. (2019). From women’s empowerment to food security: Revisiting global discourses through a cross-country analysis. Global Food Security, 23, 160–172. https://doi.org/10.1016/j.gfs.2019.05.003

Cohuet, S., Marquer, C., Shepherd, S., Captier, V., Langendorf, C., Ale, F., Phelan, K., Manzo, M. L., & Grais, R. F. (2012). Intra-household use and acceptability of Ready-to-Use-Supplementary-Foods distributed in Niger between July and December 2010. Appetite, 59(3), 698–705. https://doi.org/10.1016/j.appet.2012.07.019

Farmer, P. E., Nizeye, B., Stulac, S., & Keshavjee, S. (2006). Structural Violence and Clinical Medicine. PLOS Medicine, 3(10), e449. https://doi.org/10.1371/journal.pmed.0030449

Franklin, T. (2020). Second Uganda Nutrition Action Plan Passed. Office of the Prime Minister, Government of Uganda. https://opm.go.ug/2020/09/29/second-uganda-nutrition-action-plan-passed/

Isanaka, S., Berthé, F., Nackers, F., Tang, K., Hanson, K. E., & Grais, R. F. (2020). Feasibility of engaging caregivers in at-home surveillance of children with uncomplicated severe acute malnutrition. Maternal & Child Nutrition, 16(1), e12876. https://doi.org/10.1111/mcn.12876

Kajjura, R. B., Veldman, F. J., & Kassier, S. M. (2019). Maternal sociolJdemographic characteristics and associated complementary feeding practices of children aged 6–18 months with moderate acute malnutrition in Arua, Uganda. Journal of Human Nutrition and Dietetics, 32(3), 303–310. https://doi.org/10.1111/jhn.12643

Kapborg, I., & Berterö, C. (2002). Using an interpreter in qualitative interviews: Does it threaten validity? Nursing Inquiry, 9(1), 52–56. https://doi.org/10.1046/j.1440-1800.2002.00127.x

Kim, J. Y., Farmer, P., & Porter, M. E. (2013). Redefining global health-care delivery. The Lancet, 382(9897), 1060–1069. https://doi.org/10.1016/S0140-6736(13)61047-8

Kumeh, O. W., Fallah, M. P., Desai, I. K., Gilbert, H. N., Silverstein, J. B., Beste, S., Beste, J., Mukherjee, J. S., & Richardson, E. T. (2020). Literacy is power: Structural drivers of child malnutrition in rural Liberia. BMJ Nutrition, Prevention & Health, 3(2). https://doi.org/10.1136/bmjnph-2020-000140

Lanfer, H. L., & Reifegerste, D. (2021). Embracing challenging complexity: Exploring handwashing behavior from a combined socioecological and intersectional perspective in Sierra Leone. BMC Public Health, 21(1), 1857. https://doi.org/10.1186/s12889-021-11923-1

Liu, F., & Maitlis, S. (2010). Non-participant observation. In A. J. Mills, G. Durepos, & E. Wiebe (Eds.), Encyclopedia of case study research. / Volume 2. SAGE. http://www.worldcat.org/oclc/811140520

Mackay, H. (2019). A feminist geographic analysis of perceptions of food and health in Ugandan cities. Gender, Place & Culture, 26(11), 1519–1543. https://doi.org/10.1080/0966369X.2018.1555148

Mahmudiono, T., Segalita, C., & Rosenkranz, R. R. (2019). Socio-Ecological Model of Correlates of Double Burden of Malnutrition in Developing Countries: A Narrative Review. International Journal of Environmental Research and Public Health, 16(19), 3730. https://doi.org/10.3390/ijerph16193730

Majamanda, J., Maureen, D., Munkhondia, T. M., & Carrier, J. (2014). The Effectiveness of Community-Based Nutrition Education on the Nutrition Status of Under-five Children in Developing Countries. A Systematic Review. Malawi Medical Journal, 26(4), 115–118. https://doi.org/10.4314/mmj.v26i4

Mbogo, A. M., Van Niekerk, E., Ogada, I., & Schubl, C. (2020). Carers’ knowledge of treatment of severe acute malnutrition at Dadaab refugee complex, Kenya: A prospective cohort study. South African Journal of Child Health, 14(3), 110. https://doi.org/10.7196/SAJCH.2020.v14i3.1567

Mejia-Mantilla, C. (2020). Poverty & Equity Brief—Sub-Saharan Africa: Uganda. World Bank Group - Poverty & Equity. https://databank.worldbank.org/data/download/poverty/33EF03BB-9722-4AE2-ABC7-AA2972D68AFE/Global_POVEQ_UGA.pdf

Moore, G. F., Audrey, S., Barker, M., Bond, L., Bonell, C., Hardeman, W., Moore, L., O’Cathain, A., Tinati, T., Wight, D., & Baird, J. (2015). Process evaluation of complex interventions: Medical Research Council guidance. BMJ, 350(mar19 6), h1258–h1258. https://doi.org/10.1136/bmj.h1258

Namayengo, F. M. (2001). Substitution of milk with high-energy high-protein lactic fermented maize-bean mixture in rehabilitation of severely malnourished children [University of Nairobi]. http://erepository.uonbi.ac.ke/bitstream/handle/11295/20941/Namayengo_Substitution%20of%20milk%20with%20high-energy%20high-protein%20lactic%20fermented%20maize-%20bean%20mixture%20in%20rehabilitation%20of%20severely%20malnourished%20children.pdf?sequence=3

Nankinga, O., Kwagala, B., & Walakira, E. J. (2019). Maternal employment and child nutritional status in Uganda. PLoS ONE, 14(12). https://doi.org/10.1371/journal.pone.0226720

National Planning Authority. (2013). Uganda—Vision 2040 (Uganda Vision 2040, p. 136). National Planning Authority, Government of Uganda. https://www.greengrowthknowledge.org/sites/default/files/downloads/policy-database/UGANDA%29%20Vision%202040.pdf

National Planning Authority. (2020). Third National Development Plan (NDPIII) 2020/21— 2024/25 (Uganda Vision 2040, p. 341). National Planning Authority, Government of Uganda. http://www.npa.go.ug/wp-content/uploads/2020/08/NDPIII-Finale_Compressed.pdf

Nisbett, N., Gillespie, S., Haddad, L., & Harris, J. (2014). Why Worry About the Politics of Childhood Undernutrition? World Development, 64, 420–433. https://doi.org/10.1016/j.worlddev.2014.06.018

Nisbett, N., & Harris, J. (2018). Equity in Social and Development Studies Research: Insights for Nutrition. https://opendocs.ids.ac.uk/opendocs/handle/20.500.12413/13991

Nowell, L. S., Norris, J. M., White, D. E., & Moules, N. J. (2017). Thematic Analysis: Striving to Meet the Trustworthiness Criteria. International Journal of Qualitative Methods, 16(1). https://doi.org/10.1177/1609406917733847

Peter, E. S., Aliyu, S. H., & Hassan, R. S. (2019). Nutrition Assessment and Factors Influencing Malnutrition among Children under Five in Adjumani District Uganda. Journal of Advances in Medicine and Medical Research, 1–7. https://doi.org/10.9734/jammr/2019/v29i330074

Reed, S., & Kouam, C. E. (2013). Evaluation of community management of acute malnutrition (CMAM): Global Synthesis Report (UNICEF Evaluation Office). United Nations Children’s Fund. https://cupdf.com/document/evaluation-synthesis-report.html

Saldaña, J. (2013). The coding manual for qualitative researchers (2nd ed). SAGE.

Shah More, N., Waingankar, A., Ramani, S., Chanani, S., D’Souza, V., Pantvaidya, S., Fernandez, A., & Jayaraman, A. (2018). Community-Based Management of Acute Malnutrition to Reduce Wasting in Urban Informal Settlements of Mumbai, India: A Mixed-Methods Evaluation. Global Health: Science and Practice, 6(1), 103–127. https://doi.org/10.9745/GHSP-D-17-00182

Sharma, A. J., & Subramanyam, M. A. (2021). The intersectional role of paternal gender-equitable attitudes and maternal empowerment on child undernutrition in India. MedRxiv, 2021.02.03.21251084. https://doi.org/10.1101/2021.02.03.21251084

Shimpuku, Y., & Norr, K. F. (2012). Working with interpreters in cross-cultural qualitative research in the context of a developing country: Systematic literature review. Journal of Advanced Nursing, 68(8), 1692–1706. https://doi.org/10.1111/j.1365-2648.2012.05951.x

Soft Power Health. (2019). Soft Power Health Annual Report 2019 (p. 6). Soft Power Health. https://www.softpowerhealth.org/uploads/1/2/7/2/127289809/sph2019annualreport.pdf

Soft Power Health. (2020). Allan Stone Community Health Clinic. Soft Power Health. https://www.softpowerhealth.org/soft-power-health-clinic.html

Somassè, Y. E., Dramaix, M., Bahwere, P., & Donnen, P. (2016). Relapses from acute malnutrition and related factors in a community-based management programme in Burkina Faso: Relapses in a community-based management programme of acute malnutrition. Maternal & Child Nutrition, 12(4), 908–917. https://doi.org/10.1111/mcn.12197

Squires, A. (2009). Methodological challenges in cross-language qualitative research: A research review. International Journal of Nursing Studies, 46(2), 277–287. https://doi.org/10.1016/j.ijnurstu.2008.08.006

Tadesse, E., Berhane, Y., Hjern, A., Olsson, P., & Ekström, E.-C. (2015). Perceptions of usage and unintended consequences of provision of ready-to-use therapeutic food for management of severe acute child malnutrition. A qualitative study in Southern Ethiopia. Health Policy and Planning, 30(10), 1334–1341. https://doi.org/10.1093/heapol/czv003

Tadesse, E., Ekström, E.-C., & Berhane, Y. (2016). Challenges in Implementing the Integrated Community-Based Outpatient Therapeutic Program for Severely Malnourished Children in Rural Southern Ethiopia. Nutrients, 8(5), 251. https://doi.org/10.3390/nu8050251

Tesfay, F. H., Ziersch, A., Mwanri, L., & Javanparast, S. (2021). Experience of nutritional counselling in a nutritional programme in HIV care in the Tigray region of Ethiopia using the socio-ecological model. Journal of Health, Population, and Nutrition, 40(1), 34. https://doi.org/10.1186/s41043-021-00256-9

Tette, E. M. A., Sifah, E. K., Nartey, E. T., Nuro-Ameyaw, P., Tete-Donkor, P., & Biritwum, R. B. (2016). Maternal profiles and social determinants of malnutrition and the MDGs: What have we learnt? BMC Public Health, 16(1), 214. https://doi.org/10.1186/s12889-016-2853-z

Thorne, S., Kirkham, S. R., & MacDonald-Emes, J. (1997). Interpretive description: A noncategorical qualitative alternative for developing nursing knowledge. Research in Nursing & Health, 20(2), 169–177.

Thorne, S., Reimer Kirkham, S., & O’Flynn-Magee, K. (2004). The Analytic Challenge in Interpretive Description. International Journal of Qualitative Methods, 3(1).

Uddin, Md. F., Molyneux, S., Muraya, K., Hossain, Md. A., Islam, Md. A., Shahid, A. S. M. S. B., Zakayo, S. M., Njeru, R. W., Jemutai, J., Berkley, J. A., Walson, J. L., Ahmed, T., Sarma, H., & Chisti, M. J. (2021). Gender-related influences on adherence to advice and treatment-seeking guidance for infants and young children post-hospital discharge in Bangladesh. International Journal for Equity in Health, 20(1), 64. https://doi.org/10.1186/s12939-021-01404-7

Uganda Bureau of Statistics, & ICF. (2018). Uganda Demographic and Health Survey 2016. The DHS Program.

United Nations. (2021). Goal 2: Zero Hunger [United Nations]. Sustainable Development Goals. https://www.un.org/sustainabledevelopment/hunger/

USAID. (2021). Uganda: Nutrition Profile. USAID: From the American People. https://www.usaid.gov/global-health/health-areas/nutrition/countries/uganda-nutrition-profile

Ware, S. G. (2018). Perceptions and experiences of caregivers of severely malnourished children receiving inpatient care in Malawi: An exploratory study. Malawi Medical Journal, 30(3), 167. https://doi.org/10.4314/mmj.v30i3.7

World Health Organization. (2006). WHO child growth standards: Length/height-for-age, weight-for-age, weight-for-length, weight-for-height and body mass index-for-age: methods and development (No. 924154693X; p. 312). World Health Organization. https://www.who.int/publications-detail-redirect/924154693X

World Health Organization. (2020). Malnutrition. World Health Organization. https://www.who.int/news-room/fact-sheets/detail/malnutrition

Yebyo, H. G., Kendall, C., Nigusse, D., & Lemma, W. (2013). Outpatient Therapeutic Feeding Program Outcomes and Determinants in Treatment of Severe Acute Malnutrition in Tigray, Northern Ethiopia: A Retrospective Cohort Study. PLoS ONE, 8(6), e65840. https://doi.org/10.1371/journal.pone.0065840

Fikrie, A., Alemayehu, A., & Gebremedhin, S. (2019). Treatment outcomes and factors affecting time-to-recovery from severe acute malnutrition in 6–59 months old children admitted to a stabilization center in Southern Ethiopia: A retrospective cohort study. Italian Journal of Pediatrics, 45(1), 46. https://doi.org/10.1186/s13052-019-0642-x

Humbwavali, J. B., Giugliani, C., Nunes, L. N., Dalcastagnê, S. V., & Duncan, B. B. (2019). Malnutrition and its associated factors: A cross-sectional study with children under 2 years in a suburban area in Angola. BMC Public Health, 19(1), 220. https://doi.org/10.1186/s12889-019-6543-5

Nisbett, N., & Harris, J. (2018). Equity in Social and Development Studies Research: Insights for Nutrition. https://opendocs.ids.ac.uk/opendocs/handle/20500.12413/13991

Saldaña, J. (2013). The coding manual for qualitative researchers (2nd ed). SAGE.

Stobaugh, H. C., Rogers, B. L., Webb, P., Rosenberg, I. H., Thakwalakwa, C., Maleta, K. M., Trehan, I., & Manary, M. J. (2018). Household-level factors associated with relapse following discharge from treatment for moderate acute malnutrition. British Journal of Nutrition, 119(9), 1039–1046. https://doi.org/10.1017/S0007114518000363

Uddin, Md. F., Molyneux, S., Muraya, K., Hossain, Md. A., Islam, Md. A., Shahid, A. S. M. S. B., Zakayo, S. M., Njeru, R. W., Jemutai, J., Berkley, J. A., Walson, J. L., Ahmed, T., Sarma, H., & Chisti, M. J. (2021). Gender-related influences on adherence to advice and treatment-seeking guidance for infants and young children post-hospital discharge in Bangladesh [Preprint]. In Review. https://doi.org/10.21203/rs.3.rs-60106/v3

